# Advancing our understanding of genetic risk factors and potential personalized strategies in pelvic organ prolapse: largest GWAS to date reveals 19 novel associated loci

**DOI:** 10.1101/2021.07.08.21260068

**Authors:** Natàlia Pujol-Gualdo, Kristi Läll, Maarja Lepamets, Estonian Biobank Research Team, Henna-Riikka Rossi, Riikka K Arffman, Terhi T Piltonen, Reedik Mägi, Triin Laisk

## Abstract

**Objectives:** To identify the genetic determinants of pelvic organ prolapse (POP) and assess the predictive ability of polygenic risk scores (PRS) alone or in combination with clinical risk factors.

**Design:** Meta-analysis of genome-wide association studies (GWAS) and PRS construction and validation.

**Setting:** GWAS summary statistics from three European datasets and individual-level data from Estonian Biobank, including phenotype questionnaire and measurement panel, together with follow-up data from linkage with national health-related registries.

**Participants:** A total of 28,086 women with POP and 546,321 controls of European ancestry. Genetic risk scores were derived from a dataset of 20,118 cases and 427,426 controls of European ancestry and validated in a target dataset of 7,896 cases and 118,895 controls. Cases were defined using ICD codes and classical risk factors were derived from questionnaire data and ICD10 codes.

**Results:** The identified novel loci reinforce the role of connective tissue abnormalities, urogenital tract development and point towards association with a range of cardiometabolic traits. A novel PRS combining 3,242,959 variants demonstrated that women in the top 5% have 1.63 (95% CI: 1.37 to 1.93) times the hazard of developing POP compared to the rest of the women. When analyzing PRS in incident POP, it showed similar predictive ability (Harrell C-statistic 0.583, sd=0.007) than five established clinical risk factors (number of children, body mass index (BMI), ever smoked, constipation and asthma) combined (Harrell C-statistic 0.588, sd=0.007) and demonstrated its incremental value in combination with these (Harrell C-statistic 0.630, sd=0.007).

**Conclusions:** The largest GWAS meta-analysis in POP to date identified 26 genetic loci which establish links between POP and connective tissue abnormalities, urogenital development and cardiometabolic health. We present a PRS for POP which provides the first potential tool for preventive strategies and early detection of higher risk susceptibility to POP including genetic risk factors.

## Introduction

Pelvic organ prolapse (POP) is characterized by a descent of pelvic organs into the vaginal cavity(1). It affects around 40% of women after menopause(2–4) and the lifetime risk of gynecological surgery for POP is up to 19% in the general female population(5). The main symptoms include a bothersome sense of vaginal bulge, urinary, bowel and/or sexual dysfunction, which substantially affects a woman’s quality of life(6,7). The most common risk factors are age, number of offspring, operative vaginal delivery and BMI(8–10). However, despite its health and economic impact, the etiology of this complex disorder remains poorly understood and there is a lack of evidence for early detection of women who are at risk of developing POP.

POP is a multifactorial disorder and genetic factors have been estimated to explain 43% of the variation in POP risk in a twin study(11). A recent genome-wide association study (GWAS) of Icelandic and UK Biobank (UKBB) cohorts reported seven loci that associate with POP and point to a role of connective tissue metabolism and estrogen exposure in its etiology(12). Nevertheless, increasing the sample size is likely to boost the power for detection of more common risk variants, thus advancing our understanding of genetic risk factors underlying POP.

Polygenic risk scores (PRS), created based on GWAS results, can show the cumulative contribution of genetic factors towards disease risk. Therefore, PRS have been utilized as a tool to stratify individuals into different risk groups in complex diseases(13). However, PRS have never been used as a tool for predicting POP development. In contrast to physical examinations, which remain to be the gold-standard for POP assessment, PRS could serve as a tool to identify women with higher genetic risk of POP and tailor individualized preventive strategies even decades before symptomatic POP appears.

Given this background, we present the largest GWAS in POP to date and further integrate it with additional data layers, such as gene expression, to propose potential causal genes in associated loci. Moreover, for the first time, we aim to construct and validate the predictive ability of PRS alone or in combination with classical risk factors, a tool that might favor preventive and personalized strategies in the future.

## Methods

### Study cohorts

Our analyses included a total of 28,086 women with POP and 546,321 controls of European ancestry from three different studies: summary level statistics from an Icelandic and UKBB GWAS meta-analysis(12) (IceUK, 15,010 cases and 340,734 female controls), and FinnGen R3 (5,518 cases and 43,366 controls), and individual level data from the Estonian Biobank (EstBB, 7,896 cases and 118,895 controls) (Supplementary Figure 1). Cases were defined as women having POP diagnosed by ICD-10: N81, ICD-9: 618 and ICD-8: 623 upon availability. Controls were defined as individuals who did not have the respective ICD codes. Cohort and genotyping characteristics are reported in Supplementary Methods.

### GWAS meta-analysis

We conducted an inverse of variance weighted fixed-effects meta-analysis with single genomic control correction using GWAMA software (v2.2.2)(14). A total of 42,454,665 variants were included in the meta-analysis of 28,086 women with POP and 546,321 female controls. Genome-wide significance was set to p < 5 × 10^−8^. Assuming a prevalence of 5% for symptomatic POP and an overall POP prevalence of 40% in the population, total-h^2^ was estimated by single-trait LD score regression using the meta-analysis summary statistics and HapMap 3 LD-scores(15,16) and converted to the liability scale.

### Gene prioritization criteria

In order to move from genetic variants to plausible candidate genes, we used the following criteria. We first selected genes which showed evidence for regulatory effect of associated variants (based on gene expression or chromatin interaction data), implemented in FUMA platform v1.3.6a(17). Then, we prioritised candidate genes considering three main evidence levels: 1) distance from the association signal; 2) whether the lead signal is in high LD (r2>0.6) with a coding variant in any of the nearby genes; 3) whether the GWAS signal colocalises with a variant that affects gene expression; 4) finally, we utilized the Mouse Genome Database(18) (http://www.informatics.jax.org) to evaluate the effect of candidate genes in relevant mouse models.

### Colocalization analyses

Colocalization analyses were conducted using COLOC (v.3.2.1) R package(19) and GWAS meta-analysis effect sizes and their variances. In the analysis we compared our significant GWAS loci to all GTEXv8 and eQTL Catalogue (https://www.ebi.ac.uk/eqtl/)(20) associations (excluding Lepik et al. 2017(21) and Kasela et al. 2017(22) due to sample overlap) within 1Mbp radius of a GWAS top signal. Prior probabilities were set to p1=1e-4, p2=1e-4 and p12=5e-6. Two signals were considered to colocalize if the posterior probability for a shared causal variant was 0.8 or higher.

### Gene-Set Analysis and Tissue / Cell-Type expression analyses

Gene-set analysis and tissue expression analysis were performed using MAGMA v1.08(23) implemented in FUMA v1.3.6a(17) and DEPICT(24), implemented in Complex-Traits Genetics Virtual Lab (CTG-VL 0.4-beta)(25).

In MAGMA v1.08, gene sets were obtained from Msigdb v7.0 for “Curated gene sets” and “GO terms”. A total of 15,485 gene terms were queried. Tissue expression analysis was performed for 53 specific tissue types using MAGMA. DEPICT is an integrative tool that based on predicted gene functions highlights enriched pathways and identifies tissues/cell types where genes from associated loci are highly expressed.

### Genetic correlation

The LDSC method and GWAS-MA summary statistics were used for testing genetic correlations(15) between POP, and data available for 516 traits in LD-Hub v1.9.3 (http://ldsc.broadinstitute.org) including traits from the following categories: lipids, smoking behaviour, anthropometric, reproduction, cardiometabolic, and a range of traits from UKBB(26,27). We accounted for multiple testing using a Bonferroni correction for 516 tests (0.05/516=8.91 × 10^−5^).

### Phenome-wide associations

Pleiotropy was assessed comparing phenotype associations for the GWAS lead variants in two databases: PhenoScanner v2(28) using the *phenoscanner* (v1.0) R package (https://github.com/phenoscanner/phenoscanner), and GWAS Catalog (e96_r2019-09-24) implemented in FUMA(17). GWAS catalog look-up also included variants in high LD with lead variants (r2>0.6). For visualization of results, a heatmap was created using the *pheatmap* library in R 3.6.1. and a modified script from (https://github.com/LappalainenLab/spiromics-covid19-eqtl/blob/master/eqtl/summary_phenoscanner_lookup.Rmd). More details on results processing are found in Supplementary Methods.

### Derivation of PRS for POP

In brief, PRS analysis requires two types of data: 1) base: summary statistics of genotype-phenotype associations at genetic variants genome-wide, and 2) target: genotypes and phenotype in individuals of an independent sample(29). We constructed a POP PRS based on the summary statistics of the meta-analyses including IceUK and FinnGen, with 20,118 cases and 427,426 controls of European ancestry, leaving out EstBB as an independent target dataset.

In short, each PRS was computed for each woman in the EstBB (N=126,791) by summing the product of the allele weighting and the allele dosage across the selected SNPs. We empirically evaluated a total of 19 different versions of PRS, implementing two different methodologies: PRSice2 (v2.3.3)(30) and LDPred(31), which use a clumping and thresholding and linkage-disequilibrium SNP-reweighting approach, respectively. Whilst PRSice2 automatically calculates and applies the PRS, in the case of LDPred, STEROID (v0.1.1) tool was used for calculating PRS for all EstBB participants (https://genomics.ut.ee/en/tools/steroid). Details on PRS calculations are reported in Supplementary Methods.

### Criteria for discovery and validation set definition

First, we divided the target dataset from EstBB into a discovery and validation dataset, according to their prevalent or incident status. The discovery dataset included 5,379 prevalent cases and 21,516 controls (4 controls per case). Since controls were defined as women who did not develop pelvic organ prolapse during follow-up, they tended to be younger than prevalent cases. In the discovery set, we tested all 19 PRS versions and selected the best PRS version for further analyses (Supplementary Figure 1).

The validation set included 2,517 incident cases and 96,139 controls, and in this set we tested the predictive ability of PRS (Supplementary Figure 1). The validation set was further filtered to a validation subset, where only incident cases and controls, which presented little or no missing data of clinical risk factors data in EstBB were kept. This included a total of 2,104 incident cases and 24,780 controls, where scores were tested alone or in combination with clinical variables (Supplementary Figure 1).

### Selection of best PRS model

The discovery set was used in the initial analyses in order to select the best predicting PRS version through a logistic regression model adjusted for age, age squared, first 10 principal components and batch effects. The model that offered the smallest p-value towards case – control discrimination was selected for further analyses.

### Predictive ability of PRS and classical risk factors

We standardised the best PRS version and also categorized it into different percentiles (<5%, 5%-15%, 15%-25%, 25%-50%, 50%-75%, 75%-85%, 85%-95%, >95%). Cox proportional hazard models were used to estimate the Hazard Ratios (HR) corresponding to one standard deviation (SD) of the continuous PRS for the validation dataset. Harrell’s C-statistic was used to characterize the discriminative ability of each PRS. Cumulative incidence estimates were computed using Kaplan-Meier method. While comparing different PRS groups with each other, age was used as a timescale to properly account for left-truncation in the data.

Next, we use the validation subset to assess the predictive ability of PRS and five clinical risk factors (number of children, BMI, ever smoked, asthma and constipation) alone or in combination, and clinical risk factors together with PRS. Information on the number of children, BMI and smoking were extracted from questionnaire data, whereas ICD10 codes J45 and K59.0 were used for asthma and constipation, respectively.

## Results

As shown in Table 1 and Figure 1, 26 loci, with 30 independently significant lead signals, were associated with POP after meta-analysis. From these, 19 loci were novel findings and seven were reported previously(12). All lead variants were present in at least two out of the three datasets analyzed and were common variants (EAF>0.05) except for one replicated (EAF=0.01, rs72624976, p=1.14 × 10^−9^) and one novel association (rs72839768, EAF=0.02, p=4.66 × 10^−9^). The effect sizes of the lead variants were modest (odds ratios ranging from 0.84 to 1.19), which is consistent with GWAS findings for other complex diseases(32). There was no evidence of excessive genomic inflation (λ=1.054) in the GWAS meta-analysis, suggesting minimal bias due to population stratification, genotyping artefacts, and/or cryptic family relationships (Figure 2). SNP heritability in the meta-analysis was estimated to be 9.4% and 17.3%, considering POP prevalence of 5% (symptomatic) and 40% (overall), respectively.

**Table 1.**
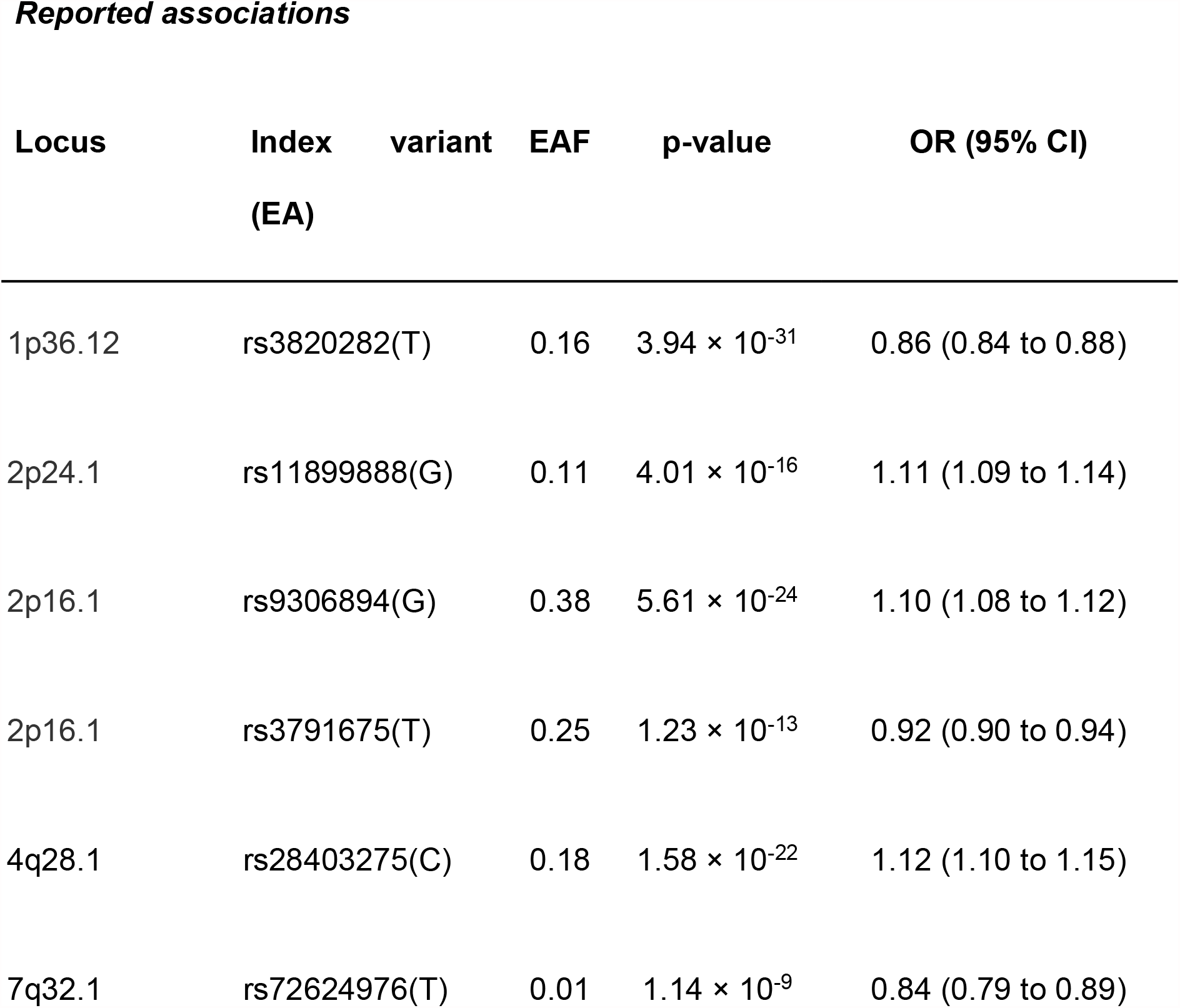
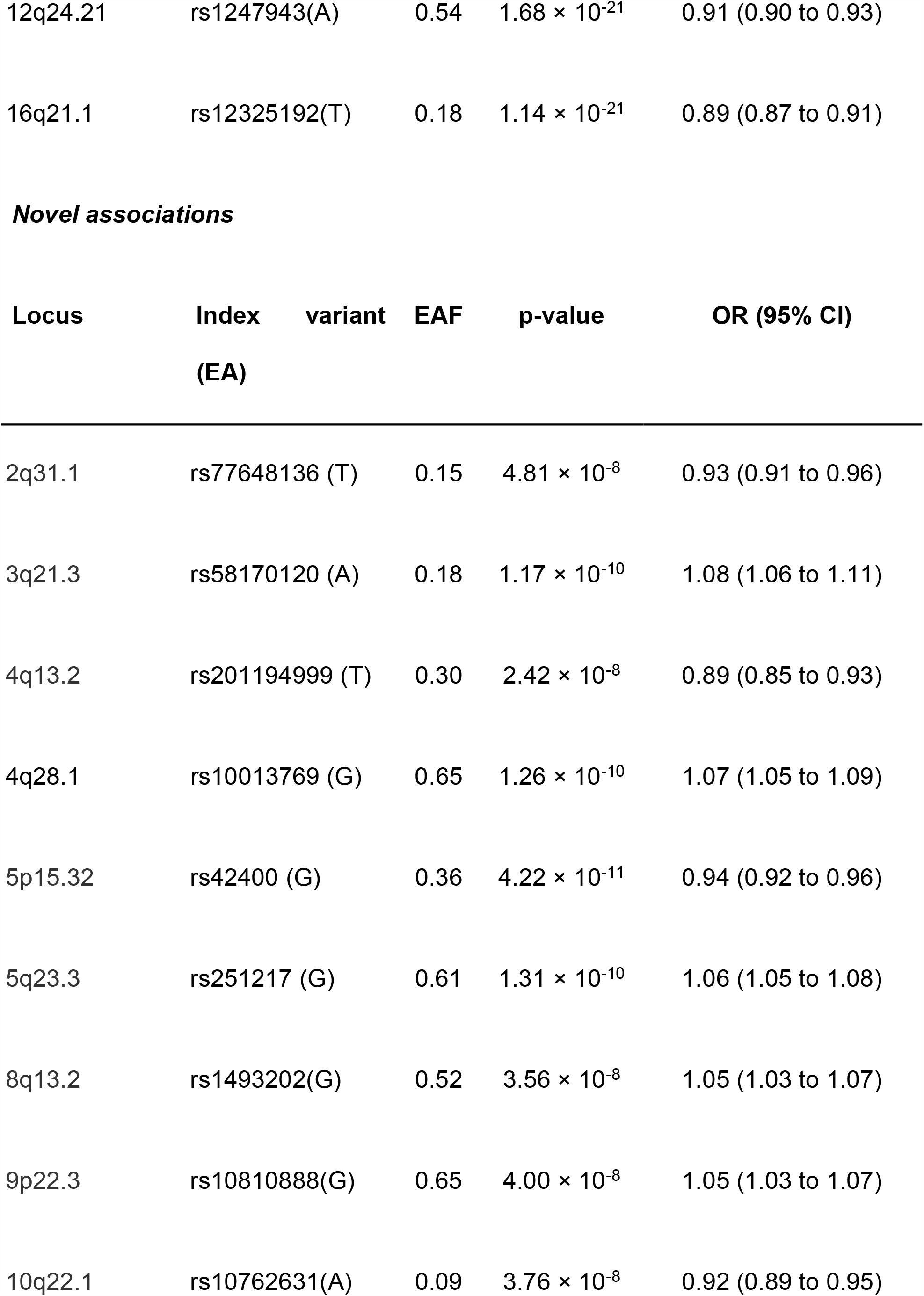
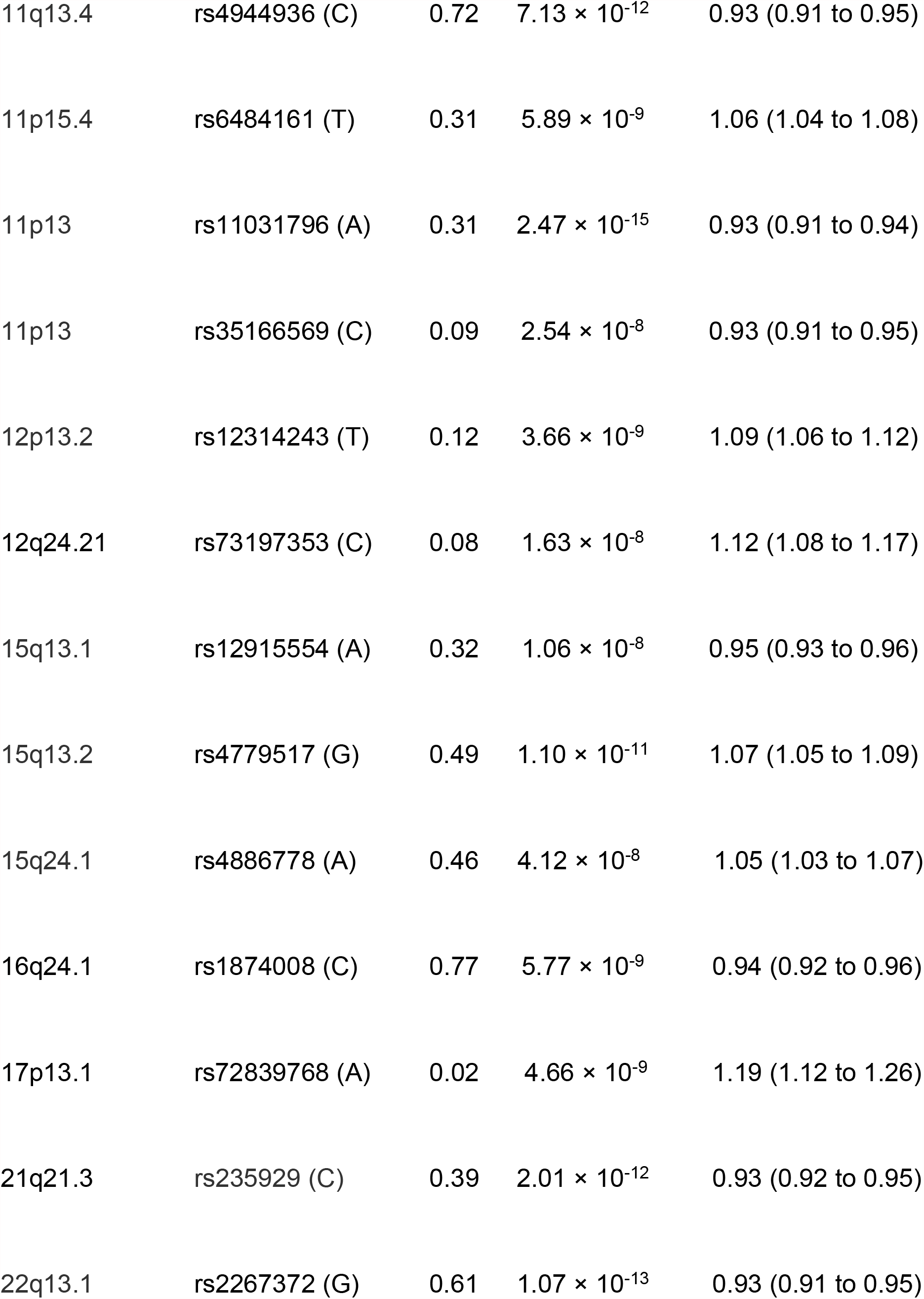
Results for the genome-wide significant index variants in the 26 loci associated with POP identified in the GWAS meta-analysis of 28,086 women with POP and 546,321 female controls.

**Figure 1.**
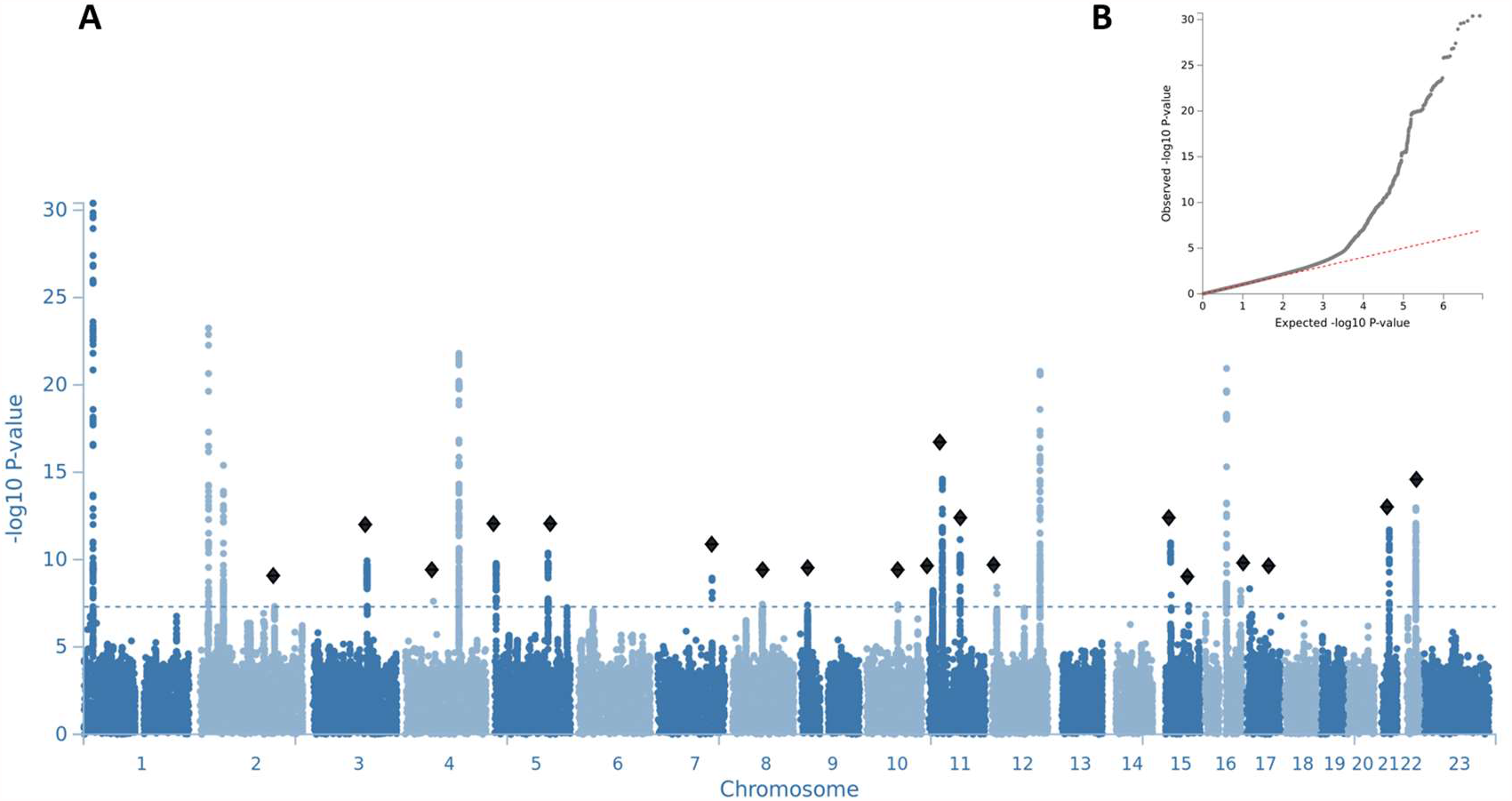
A) Manhattan plot for GWAS meta-analysis in pelvic organ prolapse and B) QQ plot. The novel candidates are highlighted as a black diamond. The y axis represents –log(two-sided P values) for association of variants with POP, from meta-analysis using an inverse-variance weighted fixed effects model. The horizontal dashed line represents the threshold for genome-wide significance.

**Figure 2.**
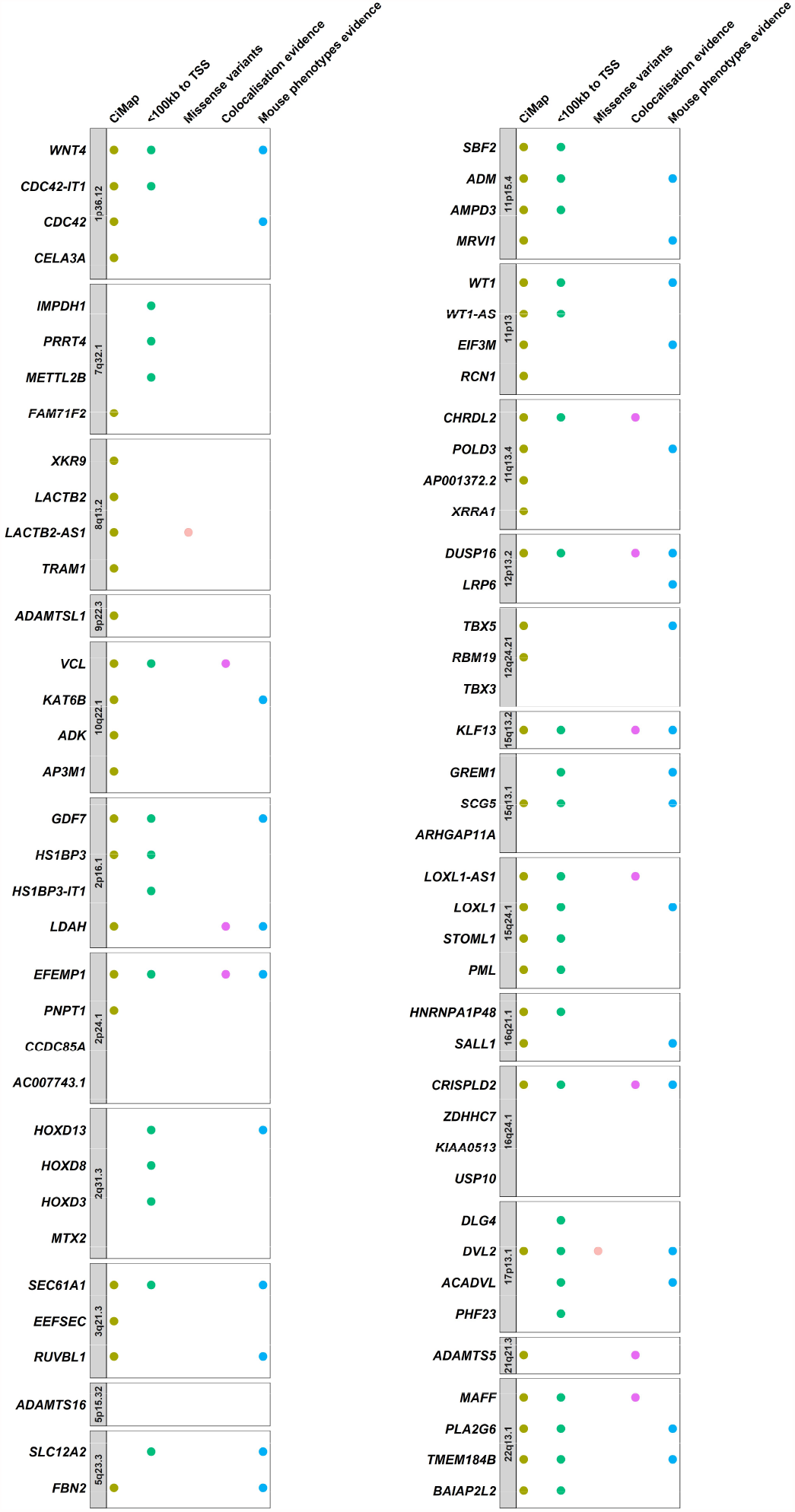
Evidence for pelvic organ prolapse GWAS meta-analysis candidate gene mapping. Genes which showed evidence for regulatory effect of associated variants (based on gene expression, which correspond to all genes listed in the figure, or also chromatin interaction data shown as yellow dots) were selected. Then, we prioritised candidate genes considering three main evidence levels: A) genes near to the association peak were prioritised (green dots indicate the genes with transcription start site (TSS) <100kb of the lead signal); B) genes containing coding variants or in high LD (r2>0.6) with these (shown as orange dots); C) genes containing shared causal variants between genetic variants and gene expression signatures unraveled by colocalization analyses (shown as pink dots); and D) genes which affected relevant phenotypes in mouse models (shown as blue dots).

### Biological mechanisms underlying associated loci

Gene prioritisation was mainly weighted by evidence supporting either distance from gene transcription start site to association peak, eQTL colocalization analyses (posterior probability for a shared causal variant PP4>0.8) and/or genes containing exonic variation (Figure 2).

Significant colocalization signals allowed us to prioritise *EFEMP1, LDAH, VCL, CHRDL2, DUSP16, LOXL1-AS1, CRISPLD2, KLF13, ADAMTS5*, and *MAFF* as potential candidate genes on 2p16.1, 2p24.1, 10q22.1, 11q13.4, 12p13.2, 15q24.1, 15q13, 16q24.1, 21q21.3, and 22q13 respectively (Figure 2, Supplementary Table 1). Data from mouse models also supported the roles of *ACADVL* (*Acadvltm1Vje/Acadvltm1Vje*), *PLA2G6* (*Pla2g6m1Sein/Pla2g6m1Sein*) and *HOXD13* (Hoxd13tm1Ddu/Hoxd13+) in muscle fiber formation, muscle weakness and muscle hypotonia phenotypes, along with *TMEM184B* knock-out mouse models which exhibited abnormal muscle fiber formation and abnormal genitalia development. Additionally, a mouse model knock-out for *LOXL1* (*Loxl1tm1Tili/Loxl1tm1Tili*) exhibited uterus prolapse and dilated uterine cervix, while *EFEMP1* knock-outs exhibited decreased skeletal muscle weight, loose skin and abnormal urogenital development.

Based on functional impact, the lead variant in 17p13.1 (rs72839768, p=4.66 × 10^−9^) is a missense variant of the *DVL2* gene. Additionally, two non-synonymous variants in LD with the lead signals were identified, *LOH12CR1* in 12p13 (rs3751262, p=2.89 × 10^−7^, r2=0.70) and *LACTB2-AS1* in 8q13.2 (rs35863913, p=3.05 × 10^−7^, r2=0.73).

### Analysis of gene set and tissue/cell-type enrichment

Gene set analysis highlighted “Connective Tissue Development” (p=1.57 × 10^−6^), “Metanephric Nephron morphogenesis” (p=3.01 × 10^−6^), “In utero embryonic development” (p=5.49 × 10^−7^), “Abnormal embryonic tissue morphology” (p=9.43 × 10^−7^) and “Small heart” (p=9.8 × 10^−6^), Supplementary Figure 2 and Supplementary Table 2). 12 tissues were significantly enriched after correcting for multiple testing, including “Cervix/ectocervix” (p=2.61 × 10^−6^), “Uterus” (p=8.16 × 10^−5^), “Embryoid bodies” (p=3.77 × 10^−6^) and “Smooth muscle” (p=5.37 × 10^−4^; Supplementary Figure 2 and Supplementary Table 3).

### Genetic correlation

Pairwise genetic correlation with POP was estimated for 561 phenotypes using LD score regression implemented in LD-Hub(15,27). 90 phenotypes demonstrated significant genetic overlap with POP (p < 8.91 × 10^−5^) (Figure 3, Supplementary Table 4). We observed the largest positive correlation with hysterectomy (r_g_=0.59, p=3.43 × 10^−17^). POP was positively correlated with the number of children (r_g_=0.22, p=2.82 × 10^−8^), whilst age at first live birth was negatively correlated (r_g_=−0.19, p=1.42 × 10^−7^). Positive associations were observed with gastroesophageal reflux (r_g_=0.31, p=5.37 × 10^−7^), diverticular disease (r_g_=0.48, p=4.33 × 10^−16^), osteoarthritis (r_g_= 0.23, p= 4.48 × 10^−6^), hiatus hernia (r_g_=0.32, p=6.68 × 10^−5^) and abdominal and pelvic pain (r_g_=0.31, p=3.58 × 10^−7^).

**Figure 3.**
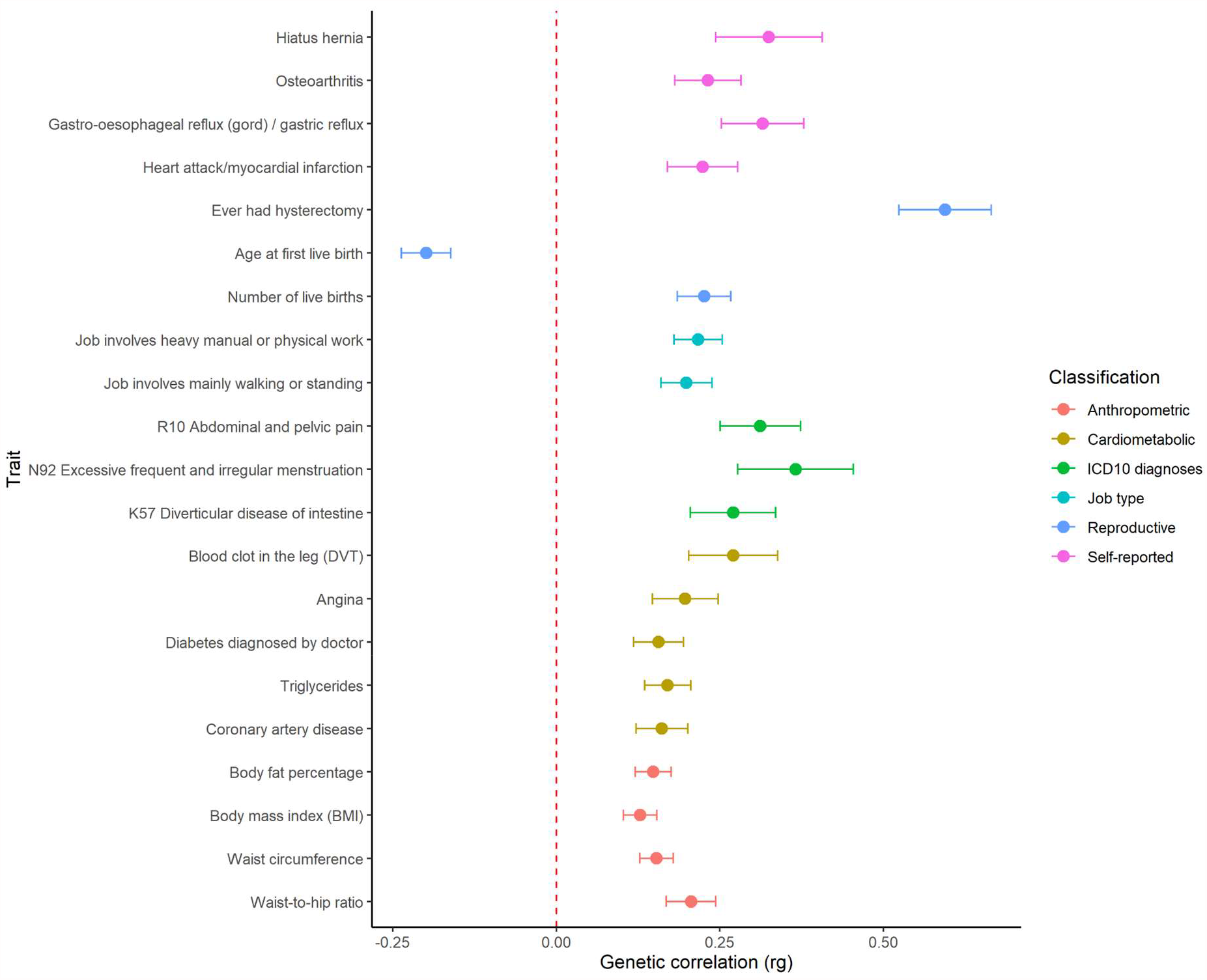
Genetic correlation analyses. Significant genetic correlations between pelvic organ prolapse (POP) and other traits reveal overlap of genetic risk factors for POP across several groups of traits (grouped by colours): anthropometric, cardiometabolic, ICD10 diagnoses, job type, reproductive traits and self-reported conditions. Center values show the estimated genetic correlation (rg), which is presented as a dot and error bars indicate 95% confidence limits.

We additionally saw positive correlations with traits such as excessive frequent and irregular menstruation (r_g_=0.36, p=3.47 × 10^−5^) and several cardiovascular phenotypes: coronary artery disease (r_g_=0.16, p=4.41 × 10^−5^), angina (r_g_=0.19, p=8.79 × 10^−5^) and myocardial infarction (r_g_= 0.22, p=3.05 × 10^−5^). Positive correlations were also observed for traits reflecting type of occupation: job involving mainly walking or standing (r_g_=0.19, p=3.7 × 10^−7^) and job involving heavy manual or physical work (r_g_=0.21, p=4.4 × 10^−9^). Genetic correlations related to obesity include significant relationships with body mass index (r_g_=0.12, p=4.73 × 10^−7^), waist-to-hip ratio (r_g_=0.20, p=4.27 × 10^−8^), triglycerides (r_g_=0.17, p=1.6 × 10^−6^) and diabetes diagnosed by doctor (r_g_=0.15, p=3.64 × 10^−5^).

### Phenome-wide associations

The phenome-wide association look-up of associated variants underlined several traits spanning abnormality of connective tissue, body measurements, cancer, cardiovascular disease, digestive system disorders, pulmonary function, reproductive health, liver disease, psychiatric disorders and other traits (Supplementary Figure 3 and 4, Supplementary Table 5 and 6).

### PRS analysis

We found that the best performing PRS consisted of 3,242,959 SNPs and was built by LDPred. This version showed an OR=1.42 (1.37 to 1.47) and p=2.59 × 10^−89^ towards the case-control discrimination in the discovery set (Supplementary Figure 5 and Supplementary Table 7).

### Predictive ability of PRS and clinical risk factors

Analyzing the best PRS with the incident POP (N=2,517 cases, 96,139 controls), a continuous PRS distribution showed the highest Harrell’s C-statistic (C-stat) of 0.616 (sd=0.006). In the validation set we observed a risk gradient within percentiles (Figure 4). Women in the top 5% of the PRS distribution had 1.63 (95% CI: 1.37 to 1.93) times the hazard of developing POP compared to the rest of the women and 1.57 (95% CI: 1.29 to 1.91) times the hazard compared to women from the average (40-60%). However, it is important to note that this is a Kaplan-Meier estimate, which does not take into account competing risks such as death before developing the disease, and thus incidence rates might be overestimated.

**Figure 4.**
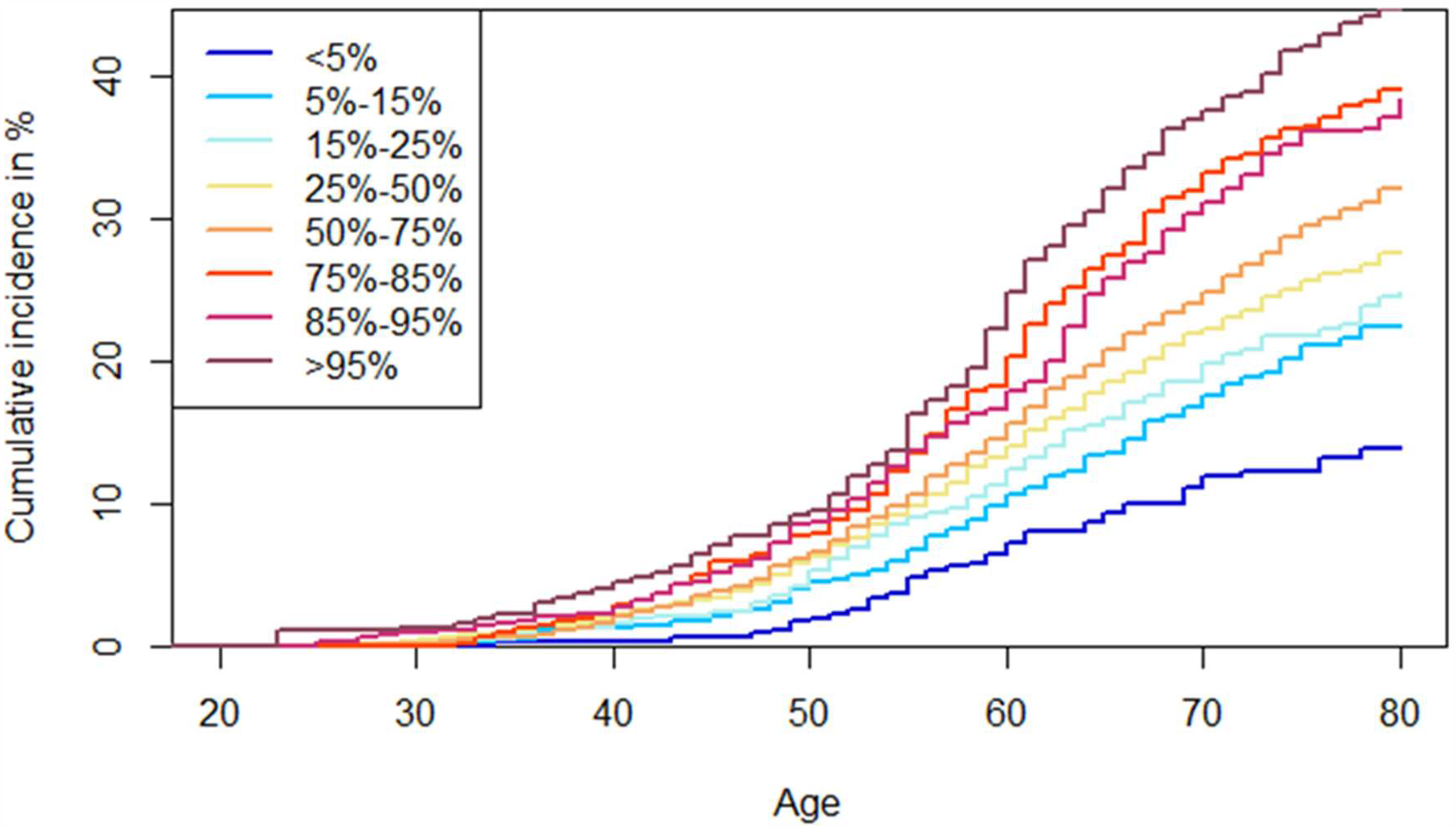
Cumulative incidence of POP in % scaled by age in the validation set of Estonian Biobank for different POP PRS percentiles. Cumulative incidence is presented separately in eight PRS categories.

In the validation subset of 2,104 cases and 24,780 controls who had clinical covariate data (Supplementary Table 8), the continuous PRS distribution showed a C-stat of 0.583 (sd= 0.007). Amongst the clinical risk factors evaluated, number of children was the best predictor (C-stat 0.563,sd=0.006), followed by ever smoked (0.534, sd=0.005), constipation (0.533,sd=0.005), BMI (0.528,sd=0.006) and asthma (0.512,sd=0.005). Adding PRS in the clinical joint model notably improved the predictive ability (0.630,sd=0.007) compared to only the clinical joint model containing the five clinical risk factors (0.588,sd=0.007) (see Figure 5 and Supplementary Table 9).

**Figure 5.**
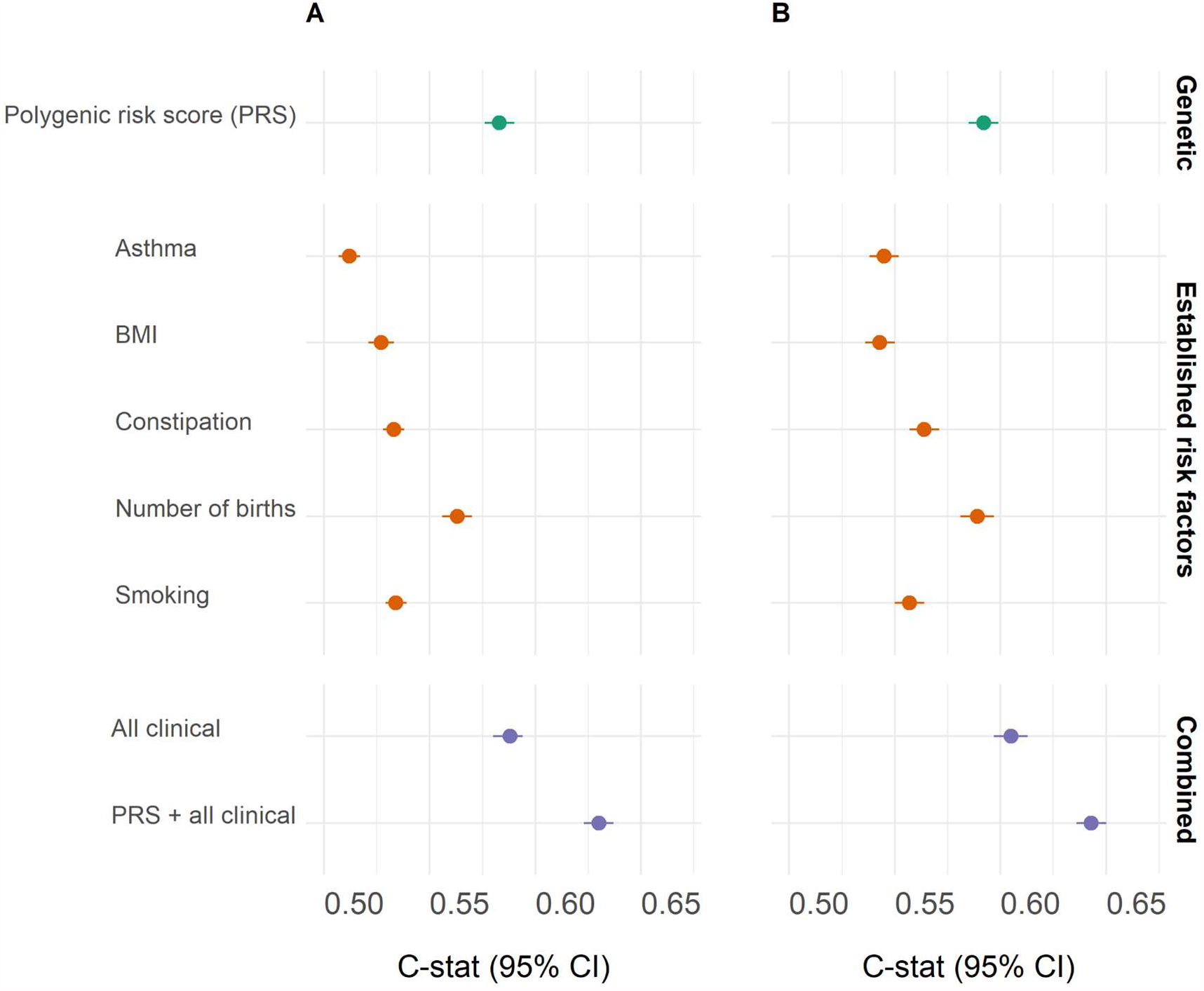
Predictive ability of PRS and clinical variables in incident status. Green dots represent polygenic risk score (PRS), orange dots represent five established risk factors and purple dots represent genetic and/or clinical combined models C-stat indexes. A) C-stat for clinical variables and PRS alone or in combination in the validation subset of Estonian Biobank. B) C-stat adjusted by batch effects and 10 first principal components in the validation subset of Estonian Biobank.

## Discussion

### Statement of principal findings

This study provides the most comprehensive analysis of genetic risk factors of POP to date, revealing many additional risk loci. We defined 26 genetic loci and used a combination of different data layers to map potential candidate genes, which provide novel insight into the biology of prolapse development, open up new avenues for further functional studies, and identify links with other health outcomes, which might have important implications for patient management and counselling. Additionally, we construct for the first time a PRS for POP, which shows similar or better predictive ability than several risk factors, such as number of children, constipation, BMI, asthma and smoking status, and we demonstrate that PRS in combination with clinical risk factors generates the best predictive model for POP. Our approach provides the first evidence to develop preventive strategies and early detection of higher risk of POP using genetic risk factors and has the potential to improve our understanding of genetic factors underlying the polygenic architecture of POP.

### Interpretation of findings and comparison to other studies

Among the genome-wide significant loci, this study supports the role of one previous reported candidate gene for pelvic organ prolapse, *LOXL1*. Liu et al. described that mice lacking the protein lysyl oxidase–like 1 (LOXL1) do not deposit normal elastic fibers in the uterine tract postpartum and develop pelvic organ prolapse(33). Subsequently, diverse mouse and human studies have reiterated its involvement with prolapse(34–37). Additionally, we further propose several candidate genes (*EFEMP1, CHRDL2, ACADVL, PLA2G6*) which reinforce the role of connective tissue molecular changes as a key process in the pathogenesis of POP(38,39). For example, *EFEMP1* encodes a fibulin and has been involved in alterations of connective tissue function as seen in inguinal hernia, pelvic organ prolapse and mouse studies(12,40–42), a link supported by a positive genetic correlation between POP and inguinal hernia in the present study. Additional genetic associations support a shared genetic background between POP and connective tissue morphology. On chromosome 21, the most plausible candidate gene was *ADAMTS5*, which encodes an ADAMTS enzyme (a disintegrin and metalloprotease with thrombospondin motifs). ADAMTS have important roles in extracellular matrix maintenance(43). A recent study described genetic associations between ADAMTS/ADAMTSL members and inguinal hernia(44), which also showed significant associations (5p13.2/*ADAMTS16* and 9p22.3/*ADAMTSL1*) in our study.

Our study also reinforces urogenital development as a key process in the pathogenesis of POP (*WNT4, DVL2, WT1, HOXD13). DVL2* and *WNT4* are components of the Wnt signaling pathway, which is important for epithelial tissue development and renewal, embryonic development of the sex organs and regulation of follicle maturation controlling steroidogenesis in the postnatal ovary(45–48). Several *in vitro* and animal studies have shown that estrogen has positive effects on the ECM(49,50) and a study described a link between hypoestrogenism and deterioration of the ECM and concomitant POP(51), indicating the WNT4 pathway may have a dual role in POP development, both by regulating organogenesis and hormonal support of tissue function.

Additionally, *WT1*, another proposed candidate gene, is a transcription factor involved in urogenital system development. Recently, a single-cell transcriptome profiling study of severe anterior vaginal prolapse described activity of WT1 in fibroblasts, and genes regulated by WT1 were enriched in terms related to actin filament behavior(52). Moreover, *WT1* has also been involved in cardiac development and disease(53–55), a link that was further supported by our phenome-wide association look-up results, since this region was associated with hypertension and cardiovascular disease (Supplementary Figure 3 and 4, Supplementary Tables 5 and 6). In this line, many associations we report point towards a link between metabolic and cardiovascular health and pelvic organ prolapse (*KLF13, DUSP16, MAFF, VCL* and *LDAH)*, which was mirrored by positive genetic correlations with a range of cardiometabolic phenotypes(56–65).

Epidemiologic studies that investigate the association between cardiovascular risk factors and female pelvic organ prolapse are currently sparse. To date, two studies reported significant associations between metabolic syndrome parameters such as elevated triglycerides or reduced HDL-cholesterol and severity of POP(66,67). Additionally, a prospective cross-sectional study that included a total of 984 middle-aged Korean women showed that metabolic syndrome was significantly associated with the pelvic floor dysfunction, including pelvic organ prolapse(68). Although a few animal experiments hypothesized that microvessel damage or chronic ischemia of the pelvic floor may compromise pelvic floor function by affecting muscle tone or innervations(69–72) the role of vascular risk factors in pelvic organ prolapse remains to be characterized and further research is warranted.

Beyond unraveling shared genetic architecture with connective tissue biology, reproductive and cardiovascular traits, genetic correlation analyses supported ‘ever had hysterectomy’ as the largest positive correlated trait with POP (rg=0.59, p=3.43 × 10^−17^). This evidence might be worth considering for obstetrician-gynecologists when hysterectomy is indicated for benign causes, although an assessment of separate groups with defined causes of hysterectomy would be needed to better interpret the direction of this association.

Additionally, genetic correlation analyses highlighted a positive correlation with job types involving walking and/or standing and heavy physical work, which might have potential value to address counselling to women with higher risk to develop POP.

### Genetic risk scores

To the best of our knowledge, this study presents the first genetic risk score model for POP, which showed similar predictive ability to several risk factors and demonstrated its added value in combination with established risk factors, in line with recent evidence for other complex diseases(73). Whilst in EstBB age was substantially increased in cases (mean=58.76,sd=12.01), than controls (mean=43.86,sd=16.06), predictive ability analyses in the validation set properly accounted for age by including it as the time scale, thus comparing only women from same ages and avoiding inflation in prediction solely due to age difference between cases and controls.

The present study also assesses the predictive ability of several clinical risk factors, showing that number of children is the most predictive clinical factor. This is in line with multiple evidence which describes parity as one of the strongest risk factors for POP(9), also mirrored by a positive genetic correlation between POP and number of children observed in this study. Constipation also had high predictive value for POP, in line with a study which observed a higher risk for constipation and lower fiber intake in women with prolapse compared to controls(74). Furthermore, our study supported smoking status as a good predictor for POP. However, there is no consensus in literature between this relationship(4,75–78). Additionally, we reinforce the relationship between BMI and pelvic organ prolapse(2,9,79). Whilst some evidence linked POP severity and bronchial asthma(80), our study concludes asthma is a poor predictor, in line with recent research which suggested that asthma is not a risk factor for POP(81). Nevertheless, these observations do not rule out the possibility that these links are established after developing POP, for instance constipation has been observed in the clinic to be both a cause and a consequence of POP, and similarly other comorbidities might appear as consequences of POP development. From a clinical point of view, we suggest that modifiable risk factors such as weight reduction and preventing constipation, along with Kegel exercises for pelvic floor muscle strengthening and avoiding heavy lifting, could be considered as preventive strategies for women with higher genetic risk to develop POP.

Recently, a validated screening tool (UR-CHOICE PFD Risk Calculator) was developed to identify pregnant women who are at higher risk for POP or other pelvic floor disorders (http://riskcalc.org/UR_CHOICE/) at 12 and 20 years after delivery(82). Models were able to discriminate between women who experienced bothersome symptoms or received treatment at 12 and 20 years, respectively, for pelvic organ prolapse (concordance indices, 0.570, 0.627). However, this risk algorithm was designed for pregnant women, thus restricting its applicability only to a narrower group of women. Contrarily, genetic risk is stable, and thus evaluable throughout the lifespan. Although the limited number of incident cases hinders assessing a predictive risk algorithm including genetic and clinical risk factors, our study demonstrates the discriminative ability of five established risk factors and clearly demonstrates the potential and incremental value of PRS when added to a clinical combined model.

### Strengths and weaknesses of the study

This study presents the largest GWAS for POP, which resulted in almost a 4-fold increase in the number of loci associated with POP. One of the strengths of this study is individual level data access in EstBB, containing genetic data of around 20% of Estonian adult population including phenotype questionnaire and measurement panel, together with follow-up data from linkage with national health-related registries, which facilitated the validation of PRS and the inclusion of clinical risk factors into a joint model.

The same biobank setting limits some aspects of the study. For instance, this study included women with POP identified using specific ICD codes, which hinders analyses considering different disease stages and severity. In the same line, the biobank setting limits the possibility to assess more extensive reproductive history information such as mode of delivery or newborn anthropometric measurements.

In regard to genetic correlation analyses, it is important to note that an observed association may not lead to inferring causality and many traits which showed a positive genetic correlation with POP are likely traditional lifestyle traits that share a strong genetic correlation without a direct causal relationship. Future research should also aim to infer causal relationships between the phenotypes uncovered by genetic correlation analyses, through Mendelian randomization principles.

Overall, this GWAS meta-analysis and PRS construction focused on European ancestry populations, which challenges the generalizability of GWAS findings to non-European populations and warrants caution when extrapolating these results to other populations.

### Unanswered questions and future research

Although gene expression data was a valuable tool for mapping potential candidate genes, future larger sample size in gene expression panels might add greater power to detect more significant associations and improve disentangling tissue specific signals in eQTL datasets(19). Whilst recent evidence suggests distance to the association peak as a good predictor for a causal gene(83,84), it is important to note that complex LD patterns between association signals might eclipse more distant genes which are the true causal ones. Overall, further functional follow-up is needed to better characterize the regulatory functions of the loci uncovered and experimental work is warranted to confirm and strengthen the role of the candidate genes we propose in the identified loci. Similarly, the role of rare variants into pelvic organ prolapse remains understudied and deserves further attention(85).

Whilst our study provides the first evidence to characterize the predictive ability of a POP PRS, future cohorts with longer follow-up time and an increased number of incident POP cases would enable more precise effect estimation and assess its translational potential to the clinic. Moreover, it remains to be determined how accurate these scores are across the lifespan; whereas genetic factors are stable, lifestyle factors and thus risk susceptibility might vary throughout time. Additionally, new PRS derived from other POP GWASs would be needed in order to assess the variability in genetic risk categorization for the same target individuals, as previous evidence showed there is considerable variability between different PRS when assessing the same complex trait(86). Our joint model shows the best predictive ability to identify women in the general population who have high risk of pelvic organ prolapse and open up novel perspectives to change screening management in the future. Whilst these are exciting opportunities, novel clinical communication and infrastructures are required in order to ethically assess the information derived from prediction models combining genetic and clinical risk data, transforming its potential usefulness into an actual benefit for women with pelvic organ prolapse.

## Data Availability

Full meta-analysis summary statistics will be made available upon publication. Icelandic and UKBB summary statistics can be accessed from http://www.decode.com/summarydata and FinnGen summary statistics can be downloaded from the browser http://r3.finngen.fi

http://r3.finngen.fi

https://gtexportal.org/home/

https://www.ebi.ac.uk/eqtl/

http://ldsc.broadinstitute.org/

http://www.informatics.jax.org/

## Web resources

FinnGen Freeze 3 PheWeb: http://r3.finngen.fi; GTEx Portal: https://gtexportal.org/home/; eQTL Catalogue: https://www.ebi.ac.uk/eqtl/; LD-Hub: http://ldsc.broadinstitute.org/; Mouse Genome Database: http://www.informatics.jax.org/;

## Acknowledgements

NPG was supported by MATER Marie Sklodowska-Curie which received funding from the European Union’s Horizon 2020 research and innovation program under grant agreement No. 813707. KL, TL, ML and RM are supported by the Estonian Research Council grant PRG687. This study was supported by European Union from the Horizon 2020 grant INTERVENE. Computations were performed in the High Performance Computing Center, University of Tartu. T.L.P., R.A. and H.R. are supported by the Academy of Finland grants no 315921 and 321763 and Sigrid Juselius foundation. We want to acknowledge the participants and investigators of the Icelandic, FinnGen, UKBB and EstBB studies. The Genotype-Tissue Expression (GTEx) Project was supported by the Common Fund of the Office of the Director of the National Institutes of Health, and by NCI, NHGRI, NHLBI, NIDA, NIMH, and NINDS. The data used for the analyses described in this manuscript were obtained from the GTEx Portal on 10/05/21. The funders had no role in study design, data collection and analysis, decision to publish, or preparation of the manuscript.

## Author contributions

Data analysis: Natàlia Pujol Gualdo, Kristi Läll, Maarja Lepamets, Reedik Mägi, Triin Laisk

Original writing: Natàlia Pujol Gualdo, Triin Laisk

Clinical assessment: Terhi T Piltonen, Henna-Riikka Rossi, Riikka K Arffman

Data: Estonian Biobank Research Team

Group authorship: Estonian Biobank Research Team

Final writing and revision: all authors

