## Supplementary Methods for "Advancing our understanding of genetic risk factors and potential personalized strategies in pelvic organ prolapse: largest GWAS to date reveals 19 novel associated loci"

#### Cohort and genotyping details

Cohort and genotyping details from Icelandic and UKBB cohorts have been reported elsewhere[1].

FinnGen is a public-private partnership project combining data from Finnish biobanks and electronic health records from different registries. In this study, we used the results from the FinnGen release R3 (http://r3.finngen.fi), which includes data from 135,638 individuals and more than 1,800 disease endpoints and it is publicly available for download. FinnGen individuals have been genotyped with Illumina and Affymetrix arrays and imputed to the population-specific SISu v3 importation reference panel. Genetic association testing has been carried out with SAIGE. We downloaded the summary statistics querying the disease endpoint "Female genital prolapse", which included 5,518 individuals with the ICD10 N81 diagnosis as cases and 43,366 controls.

The Estonian Biobank (EstBB) is a population-based biobank with over 200,000 participants, currently including around 135,000 women (20% of Estonian female population). The 200K data freeze was used for the analyses described in this paper. All biobank participants have signed a broad informed consent form. Individuals with pelvic organ prolapse were identified using the ICD-10 code N81 (mean age=58.76, sd= 12.01), and all female biobank participants who did not have this diagnosis were considered as controls (mean age=43.86, sd=16.06), which included 7,968 cases and 118,895 controls. Information on ICD codes is obtained via regular linking with the national Health Insurance Fund and other relevant databases[2].

All EstBB participants were genotyped using Illumina GSAv1.0, GSAv2.0, and GSAv2.0_EST arrays at the Core Genotyping Lab of the Institute of Genomics, University of Tartu. Samples were genotyped and PLINK format files were created using Illumina GenomeStudio v2.0.4. Individuals were excluded from the analysis if their call-rate was < 95% or if their sex defined by heterozygosity of X chromosomes did not match their sex in the phenotype data. Before imputation, variants were filtered by call-rate < 95%, HWE p-value < 1e-4 (autosomal variants only), and minor allele frequency < 1%. Variant positions were updated to b37 and all variants were changed to be from the TOP strand using GSAMD-24v1-0_20011747_A1-b37.strand.RefAlt.zip files from the https://www.well.ox.ac.uk/~wrayner/strand/ webpage. Pre-phasing was conducted using Eagle v2.3 software[3] (number of conditioning haplotypes Eagle2 uses when phasing each sample was set to: --Kpbwt=20000) and imputation was done using Beagle v.28Sep18.793[4] with effective population size ne=20,000. The population specific imputation reference of 2,297 whole genome sequencing (WGS) samples was used[5]. Association analysis was carried out using SAIGE (v0.38) software to implement a mixed logistic regression model with year of birth and 10 PCs as covariates in step I.

### Processing of PheWAS look-up results

The results obtained were filtered to keep one association per variant per trait, keeping studies from newer or larger studies. Descriptions of Experimental Factor Ontology (EFO) terms and classification of EFO broad categories were obtained from the GWAS Catalog. Missing categories were added by manually searching the EMBL-EBI EFO webpage (www.ebi.ac.uk/efo/).

### POP polygenic risk score calculation

We compared two methods for calculating a polygenic risk score (PRS) for POP. Genetic variants with MAF<0.01, indels and variants with imputation score 0.8 and lower in EstBB were removed from all polygenic risk score calculations. PRSice-2 uses a "clumping and thresholding" approach to clump genetic variants in close linkage disequilibrium, such that the remaining variants are independent of each other, and includes only those variants with a GWAS association *P*-value below a given threshold, with the threshold chosen to maximize the association of the risk score with POP. We tested the following thresholds: 1, 0.3, 0.1, 0.03, 0.01, 0.003, 0.001, 0.0003 and 0.0001, with a maximum LD between them set to r2=0.2.

LDpred is a Bayesian approach that applies a continuous shrinkage model to modify effect sizes of SNPs to incorporate information on the strength of each variant's association in the GWAS and the underlying linkage disequilibrium structure[6]. To decrease the dimension of multicollinearity, SNPs were clumped with maximum LD between them set to r2=0.99. Then, 10 versions of PRSs were calculated by varying the fraction of causal SNPs on these values: Inf, 1, 0.3, 0.1, 0.03, 0.01, 0.003, 0.001, 0.0003 and 0.0001. Possible convergence issues were reported by the program for some fractions (different depending on the base study) while Gibbs sampler tried to estimate the posterior effect estimates.
