## Supplementary Figures for "Advancing our understanding of genetic risk factors and potential personalized strategies in pelvic organ prolapse: largest GWAS to date reveals 19 novel associated loci"

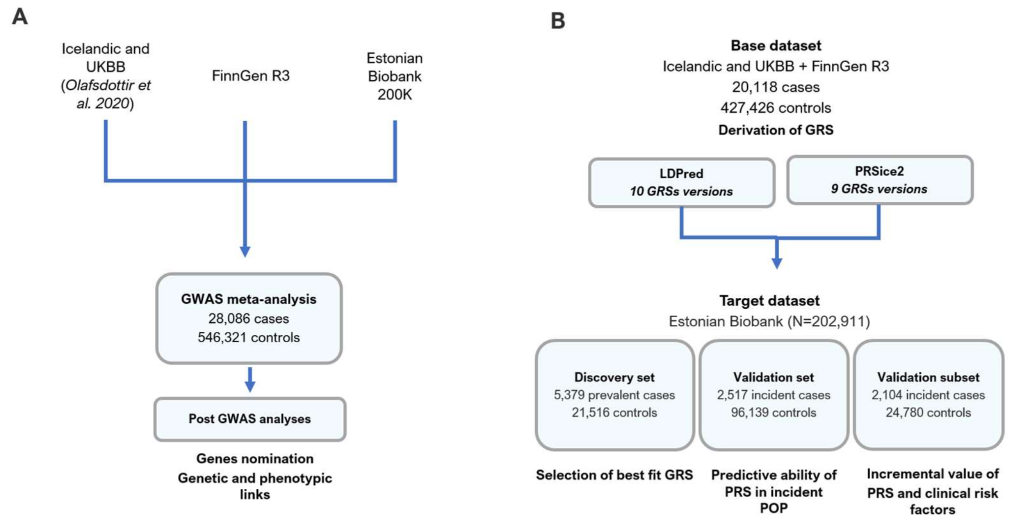


**Supplementary Figure 1. Study design.** A) Three European ancestry case-control studies were meta-analysed containing a total of 28,086 women with POP and 548,321 controls. Post-GWAS analyses guided the nomination of candidate genes and established genetic and phenotypic links. B) Derivation of PRS from base dataset and PRS construction and validation using Estonian Biobank as target dataset. Target dataset was further split into discovery and validation set and subset. UKBB: UK Biobank, GWAS; genome-wide association study, PRS: polygenic risk score.

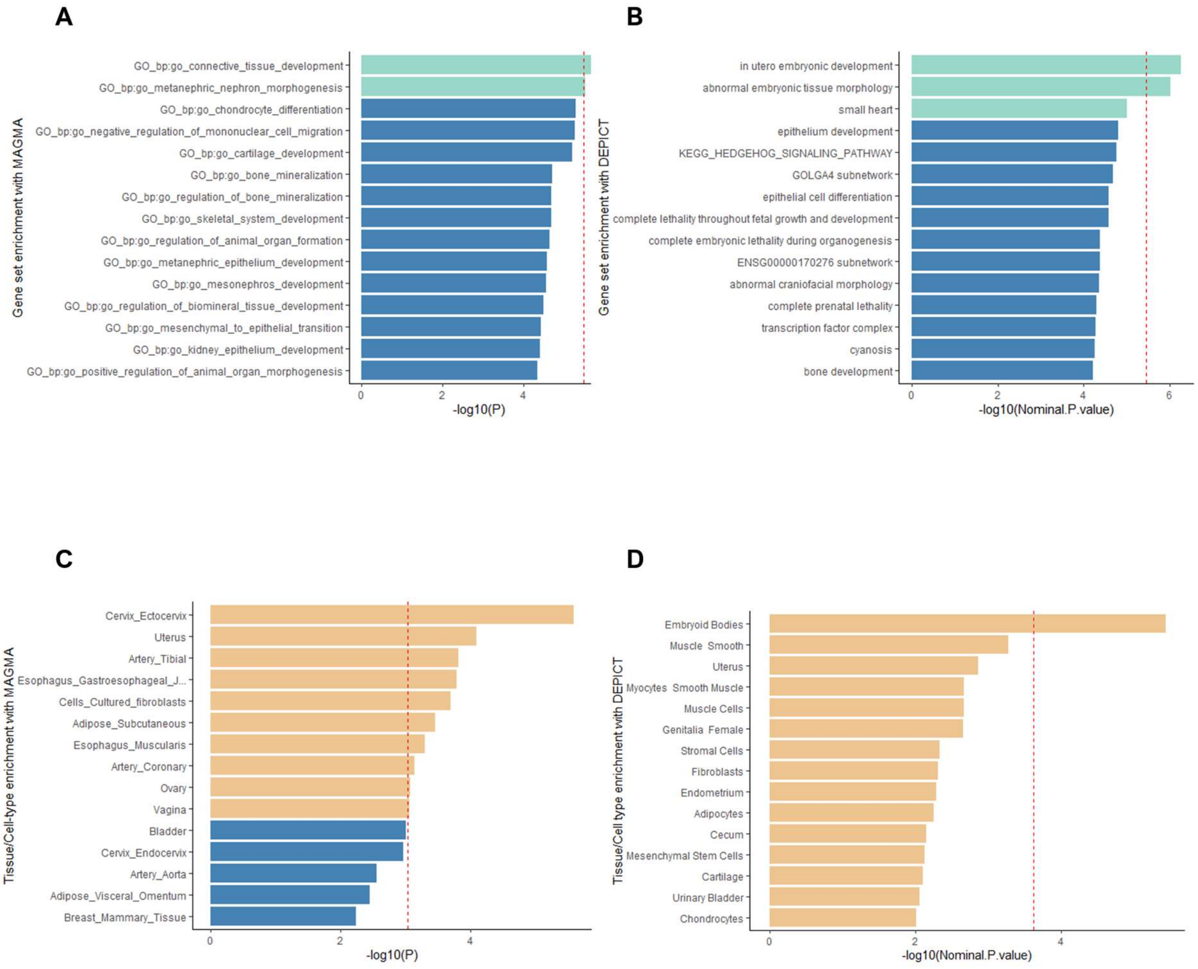


**Supplementary Figure 2. A) Gene set enrichment analyses with MAGMA.** Green bars and the red dashed line indicate Bonferroni threshold, set to p=0.05/15485=$3.22 \times 10^{-6}$. **B) Gene set enrichment analyses with DEPICT.** FDR corrected results (p<0.00001) are shown in green bars and the red dashed line indicates Bonferroni threshold set to p=0.05/14461=$3.45 \times 10^{-6}$ **C) Tissue enrichment analyses with MAGMA.** Orange bars and the red dashed line indicate Bonferroni threshold, set to p=0.05/54=0.0009**. D) Tissue/cell-type enrichment analyses with DEPICT.** FDR corrected results (p<0.00001) are shown in orange and the red dashed line indicates Bonferroni threshold set as p=0.05/209=0.0002

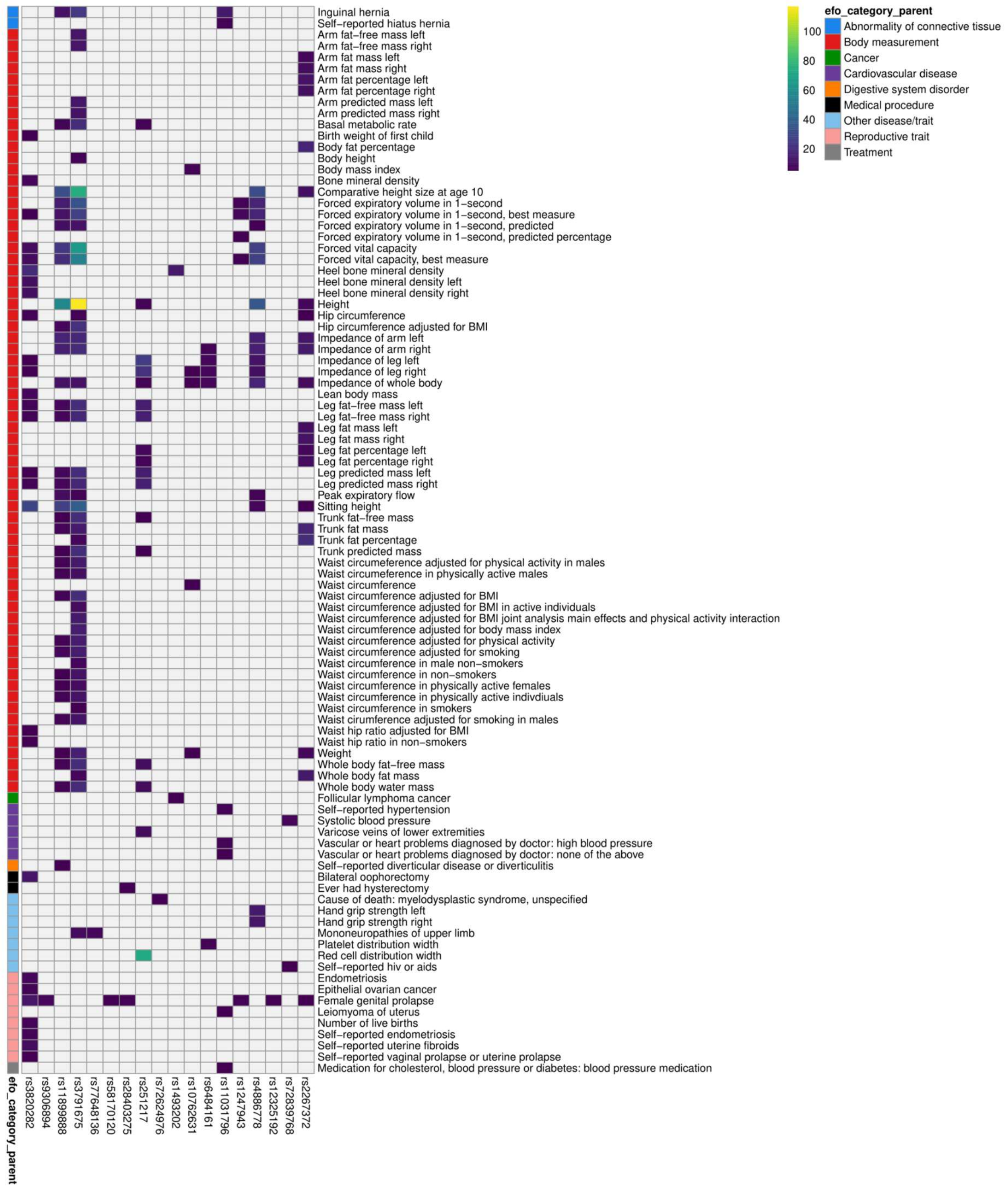


**Supplementary Figure 3. Phenoscanner look-up results.** Heatmap showing the pheWAS associations with the GWAS lead variants. Traits are grouped based on the experimental factor ontology (EFO) terms, and EFO terms with few traits grouped into one group, “Other disease/trait”. Tiles are colored by the -log10(GWAS P-value) or grey, if there was no suggestive signal obtained for the given variant and trait (GWAS P-value > 10-5).

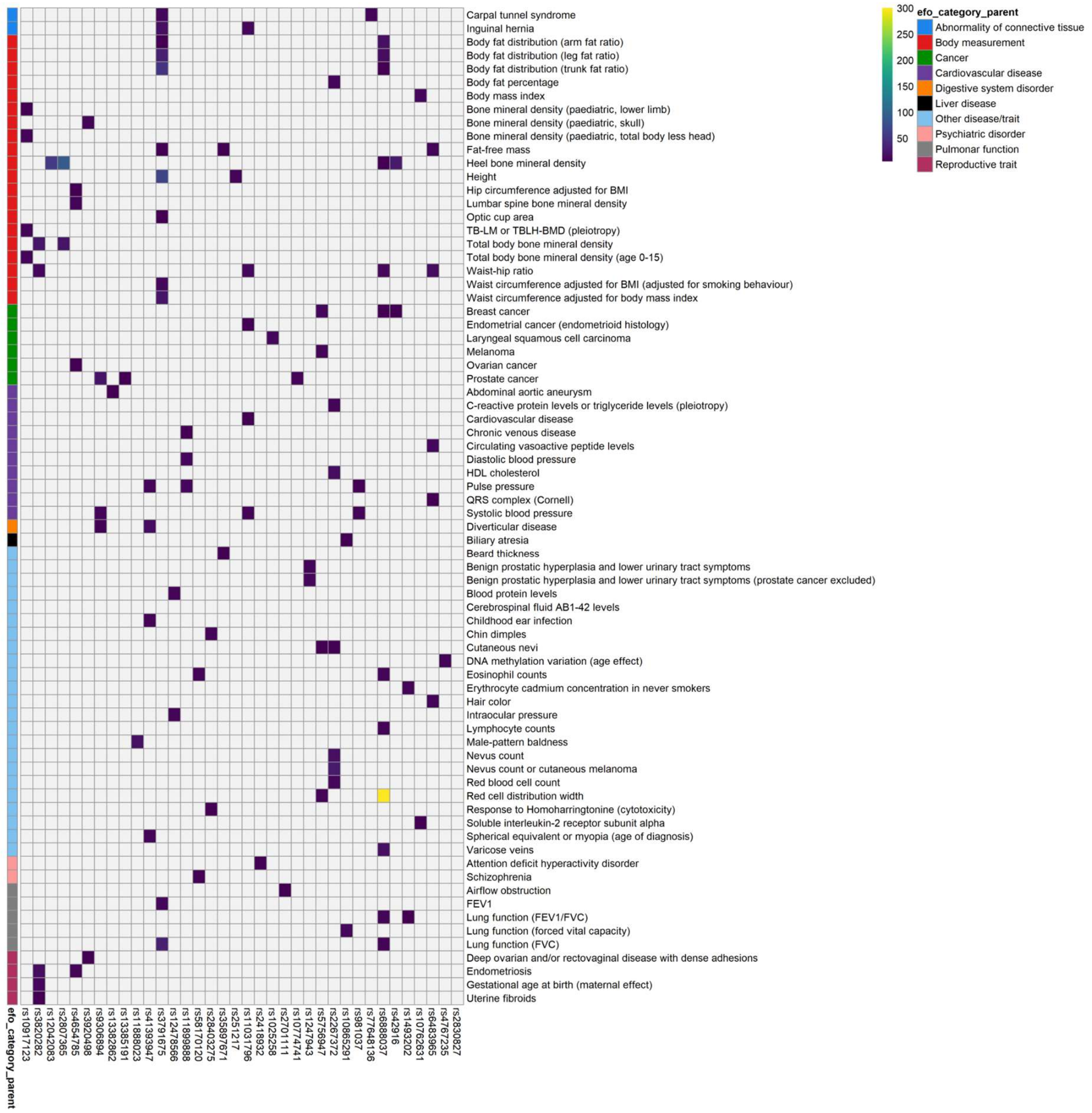


**Supplementary Figure 4. GWAS catalog results.** Heatmap showing the pheWAS associations with the GWAS lead variants and variants in high LD (r2>0.6) with these. Traits are grouped based on the experimental factor ontology (EFO) terms, and EFO terms with few traits grouped into one group, “Other disease/trait”. Tiles are colored by the -log10(GWAS P-value) or grey, if there was no suggestive signal obtained for the given variant and trait (GWAS P-value > 10-5).

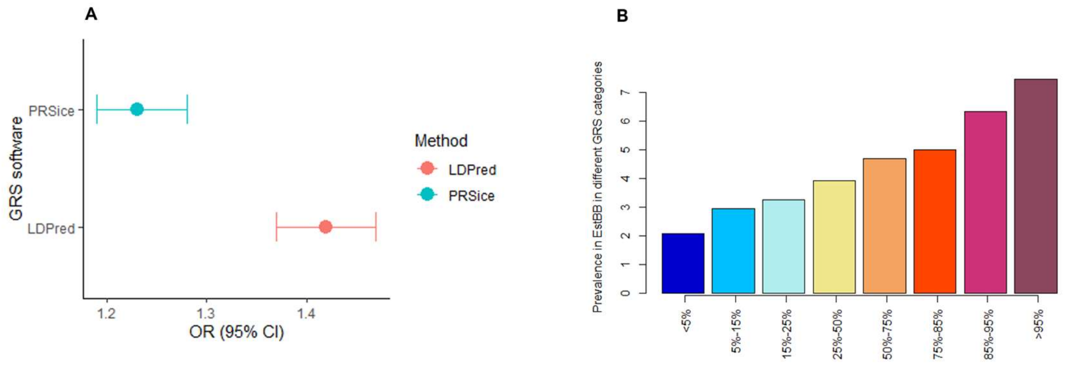


**Supplementary Figure 5. A) Comparison of best-fit PRS generated by PRSice and LDpred B) POP prevalence (%) between different PRS distributions in Estonian Biobank.**
